# Machine Learning Directed Interventions Associate with Decreased Hospitalization Rates in Hemodialysis Patients

**DOI:** 10.1101/2020.10.07.20207159

**Authors:** Sheetal Chaudhuri, Hao Han, Len Usvyat, Yue Jiao, David Sweet, Allison Vinson, Stephanie Johnstone Steinberg, Dugan Maddux, Kathleen Belmonte, Jane Brzozowski, Brad Bucci, Peter Kotanko, Yuedong Wang, Jeroen P. Kooman, Franklin W Maddux, John Larkin

## Abstract

**Background:** An integrated kidney disease company uses machine learning (ML) models that predict the 12-month risk of an outpatient hemodialysis (HD) patient having multiple hospitalizations to assist with directing personalized interdisciplinary interventions in a Dialysis Hospitalization Reduction Program (DHRP). We investigated the impact of risk directed interventions in the DHRP on clinic-wide hospitalization rates.

**Methods:** We compared the hospital admission and day rates per-patient-year (ppy) from all hemodialysis patients in 54 DHRP and 54 control clinics identified by propensity score matching at baseline in 2015 and at the end of the pilot in 2018. We also used paired T test to compare the between group difference of annual hospitalization rate and hospitalization days rates at baseline and end of the pilot.

**Results:** The between group difference in annual hospital admission and day rates was similar at baseline (2015) with a mean difference between DHRP versus control clinics of −0.008±0.09 ppy and −0.05±0.96 ppy respectively. The between group difference in hospital admission and day rates became more distinct at the end of follow up (2018) favoring DHRP clinics with the mean difference being −0.155±0.38 ppy and - 0.97±2.78 ppy respectively. A paired t-test showed the change in the between group difference in hospital admission and day rates from baseline to the end of the follow up was statistically significant (t-value=2.73, p-value<0.01) and (t-value=2.29, p-value=0.02) respectively.

**Conclusions:** These findings suggest ML model-based risk-directed interdisciplinary team interventions associate with lower hospitalization rates and hospital day rate in HD patients, compared to controls.

## Introduction

End Stage Kidney Disease (ESKD) patients frequently experience emergent complications requiring hospitalization. In the United States, ESKD patients treated by dialysis had on average 1.7 admissions during 2018 [1]. In addition to the negative consequences to the patient from the onset/exacerbation of a disease requiring inpatient care, hospitalizations are economically impactful to the healthcare system accounting for about 30% of all ESKD Medicare expenditures [1]. An array of clinical parameters associate with hospitalization events in dialysis patients, including potentially modifiable factors related to the patients’ dietary, psychosocial, and other needs [2-6]. However, classifying a dialysis patient’s risk for hospitalization, understanding the root cause of likely complications, and providing timely interventions can be challenging based on standard practices.

Most ESKD patients in the United States are treated by outpatient hemodialysis (HD) performed thrice weekly, which amasses large volumes of longitudinal clinical data in electronic medical records (EMR). This robust data collected on HD patients affords opportunities to use Artificial Intelligence (AI) methods to assist with individualized hospitalization risk classifications. AI is an overarching group of advanced analytical techniques bringing together concepts from fields such as computer science, statistics, algorithmics, machine learning (ML), information retrieval, and data science at large [7]. ML techniques are very powerful in their ability to quickly detect hidden patterns in large datasets [8] and have been reported to have the ability to assist with prediction of likely future outcomes for mortality, transplant failure, and other events in ESKD [9-13]

An integrated kidney disease company has been leveraging historic patient data with advanced analytics to direct care in quality improvement efforts at its national network of dialysis clinics. The provider has developed and operationally deployed a set of ML models that identify in-center HD patients at an increased risk for multiple all-cause hospitalizations within the next 12 months. The models were used in a pilot called the Dialysis Hospitalization Reduction Program (DHRP) that provides risk directed interdisciplinary team root cause evaluations and personalized interventions to HD patients predicted to be at risk of ≥6 hospital admissions within the next 12 months.

We investigated the impact of the DHRP on clinic-wide hospitalization rates in HD patients. Moreover, in light of other analysis supporting the use of behavioral health interventions, we explored profiles of psychosocial barriers that included patient reported outcomes for depression, sleep, and psychological stress status related to one of the DHRP interventions that was explicitly recorded in the EMR [14].

## Materials and Methods

### General Design

An integrated kidney disease company (Fresenius Medical Care, Waltham, MA, United States) developed a set of ML prediction models for classification of individual dialysis patients at risk for multiple hospitalizations within the next 12 months. These ML models have been used since 2016 to direct interdisciplinary evaluations and interventions in the DHRP quality improvement pilot being performed at select dialysis clinics among a national network (Fresenius Kidney Care, Waltham, MA, United States).

We performed an analysis to evaluate the rolling annual hospital admission and day rates per-patient-year (ppy) in clinics before (2015) and after (2016-2018) the initiation of DHRP and compared rates to matched control clinics not involved in the DHRP.

This analysis was performed under a protocol reviewed by New England Institutional Review Board (Needham Heights, MA, United States; Version 1.0 NEIRB# 17-1305247-1; Revised Version 1.1 NEIRB# 17-1344262-1) who determined this analysis of existing patient data that was de-identified by the investigator was exempt and did not require informed consent. This analysis was conducted in adherence with the Declaration of Helsinki.

### Patient Population

In this analysis, we included data from HD patients treated in a clinic participating in the DHRP quality improvement pilot implemented across the United States. For the selection of control clinics, we assessed data from the national network and matched clinics with similar attributes that were not involved in the DHRP. Control clinics were matched in a 1:1 ratio to DHRP clinics on the logit of the propensity score for the number of HD patients in the clinic, average age, percentage of male/female, percentage of white/black patients, average albumin, presence of comorbidities (congestive heart failure (CHF), diabetes, ischemic heart disease), and the hospital admission and day rates during the baseline period (2015). We excluded data from all clinics that were managed via the ESKD Seamless Care Organization (ESCO) program, irrespective of their participation in the DHRP.

### ML Models

A set of ML models were developed to classify an individual’s 12-month risk of multiple hospitalizations using historical EMR data from nearly 140,000 incenter HD patients. Overall, close to 300 different input variables were used in the ML models. The input variables included HD patient data on demographics, comorbidities, hospitalization history, treatment history, laboratories, quality of life (QOL) surveys, as well as publicly available data on social, economic, environmental, and geographical factors. (**Figure 1**). For development of the ML models, the historic data were randomly split into 50% training, 20% validation, and 30% test dataset.

**Figure 1:**
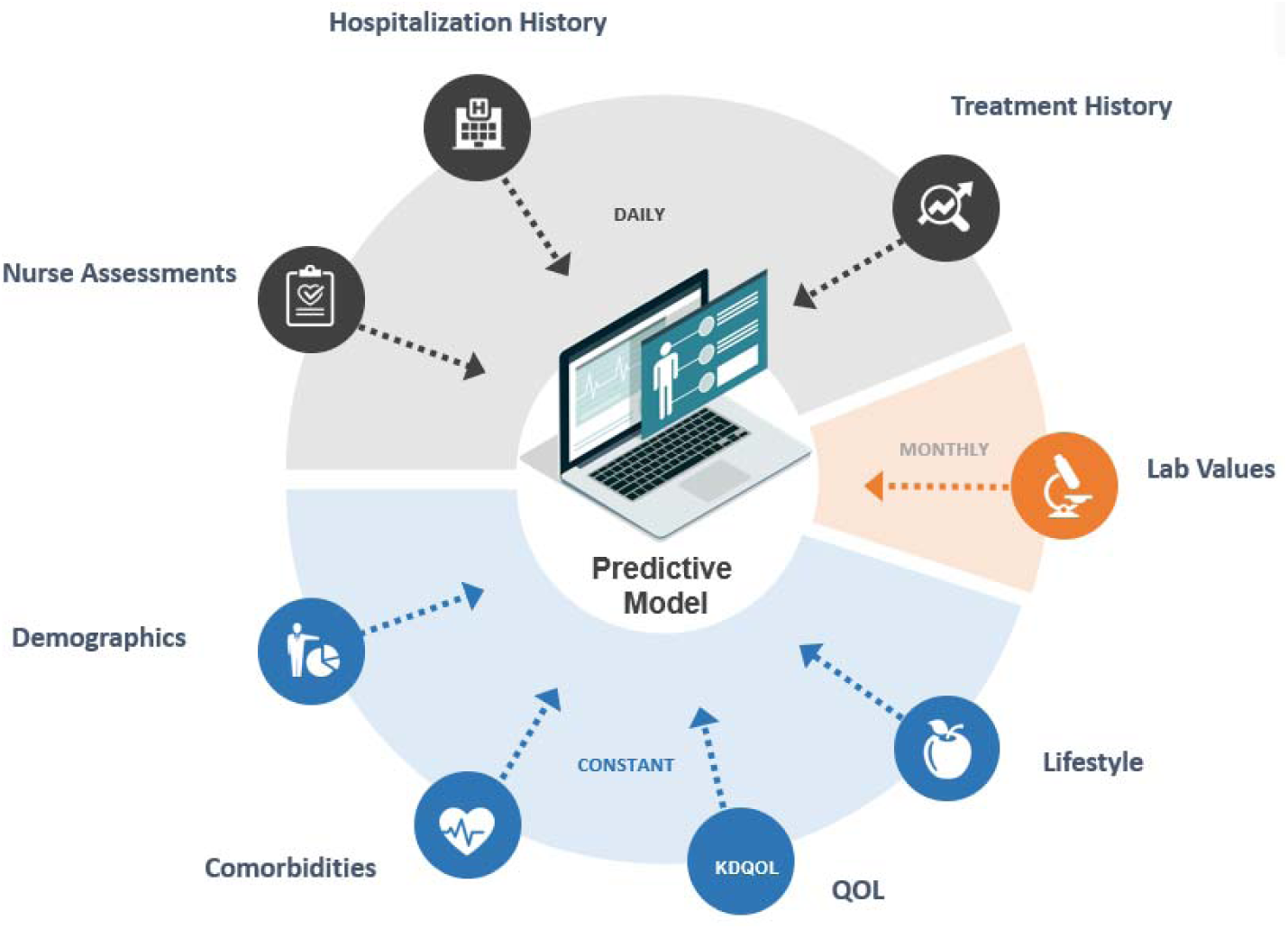
Various input sources for developing a machine learning ML model to predict patients at risk of hospital admissions in the next 12 months.

The ML models considered three binary classification tasks to predict whether an individual patient was at risk for having:

1. ≥6 hospitalizations within the following 12 months
2. ≥3 hospitalizations within the following 12 months
3. ≥1 hospitalization(s) within the following 12 months

Two models were developed for each binary classification task. One was developed for patients who were received incenter HD treatments for at least 120 days, and the other one was developed for patients with less than 120 days of incenter HD treatments. Thus, there were six models developed as shown in **Table 1**; all six models were gradient boosting models built using XGboost package [15].

**Table 1:** 6 Variations of the ML model.

| | Exposure days $\geq 120$ (established-patient models) | Exposure days $< 120$ (new-patient models) |
| --- | --- | --- |
| Risk of Hospitalization in the following 12 months $\geq 6$? | Model 1 * (AUROC: 0.813) | Model 2* (AUROC: 0.801) |
| Risk of Hospitalization in the following 12 months $\geq 3$? | Model 3 (AUROC:0.776) | Model 4 (AUROC:0.758) |
| Risk of Hospitalization in the following 12 months $\geq 1$? | Model 5 (AUROC:0.724) | Model 6 (AUROC:0.701) |
| All Area Under the Receiver Operating Curve (AUROC) was determined from the test dataset |  |  |
| *Models used for risk directed care in DHRP |  |  |

**Table 2:** Demographic and comorbidities breakdown of patients in DHRP and control clinics at baseline.

|  | DHRP | Control | Mean Std Diff |
| --- | --- | --- | --- |
| Number of Clinics | 54 | 54 |  |
| Average Number of Patients | 182 | 188 | -0.067 |
| Average Age | 61.6 | 61.0 | 0.201 |
| Male (%) | 57% | 56% | 0.113 |
| White (%) | 61% | 60% | 0.027 |
| Black (%) | 33% | 37% | -0.156 |
| CHF (%) | 21.5% | 23.5% | -0.252 |
| Diabetes (%) | 68.1% | 68.8% | -0.083 |
| Hypertension (%) | 65.5% | 70.1% | -0.368 |
| IHD (%) | 18.6% | 20.1% | -0.138 |
| Average Albumin | 3.75 | 3.74 | 0.275 |
| Hospital Admission Rate in 2015 | 1.35 | 1.35 | 0 |
| Hospital Admission Days in 2015 | 9.26 | 9.26 | 0 |
| CHF: Chronic Heart Failure, IHD: Ischemic Heart Disease |  |  |  |

For each patient, the models predicted the probability of having at least 6 hospital admissions, at least 3 hospital admissions, and at least 1 hospital admission in the following 12 months; we defined these risk classifications as high-risk, medium-high-risk, and medium-low-risk respectively. **Figure 2** shows how the risk category of each patient was designated given the predicated probabilities. For the ML model directed interventions in the DHRP, the interdisciplinary teams utilized the two high risk models that identified patients at risk of at least 6 hospital admissions in the next 12 months. The classifications from the other four models were not utilized for interventions but was used for assessing the patient progress over time.

**Figure 2:**
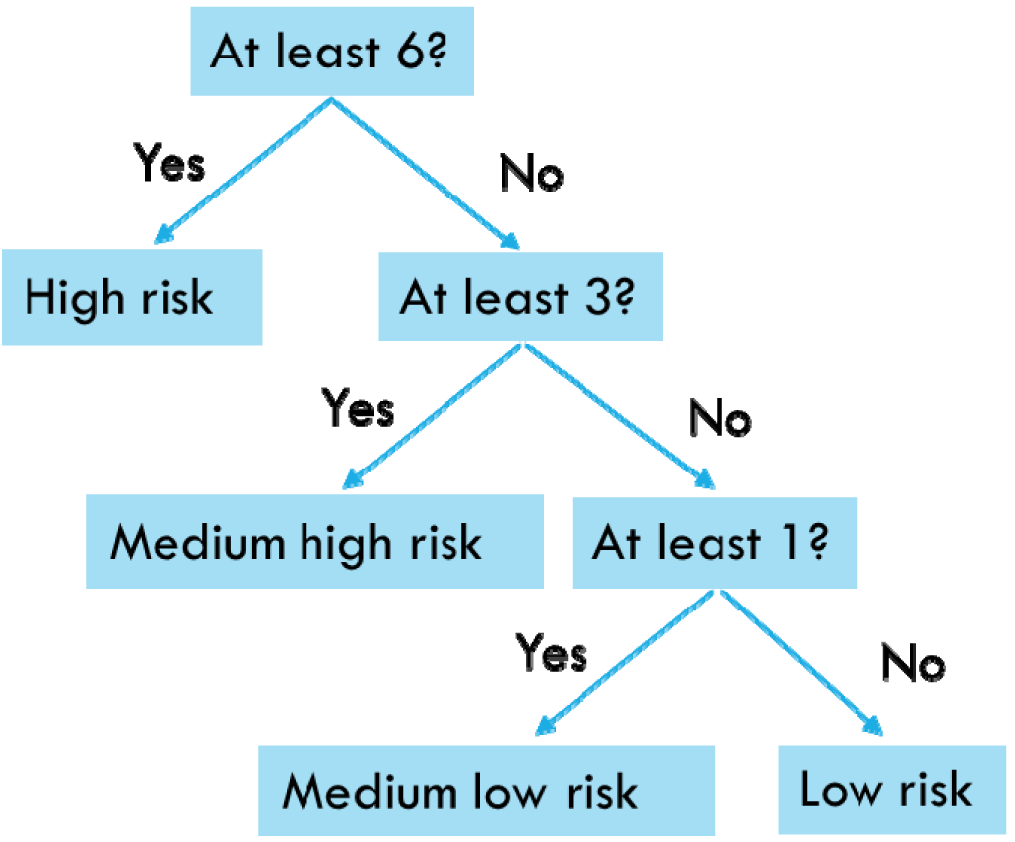
Decision tree to label High-Risk, Medium-High-Risk, Medium-Low-Risk and Low-Risk patients.

The performance of the model was evaluated using area under the receiver operating characteristic curve (AUROC) [16]. The AUROC for the 6 models was between 0.70 to 0.81 on the test data (**Table 1**).

### Interventions

Starting in 2016, DHRP clinics received a list of high-risk patients via a secured email and spreadsheet every month. The clinic manager, or a nurse assigned to lead the program, partnered with an interdisciplinary team of social workers, dieticians, and nurses to triage evaluations/interventions to high-risk patients. The interdisciplinary team performed a root cause analysis of risk for hospitalization for each patient identified as high-risk by the ML model. Each team member formulated individualized goals to address any medical instability that could lead to a hospitalization. Also, routine brief meetings were scheduled to discuss team actions taken and next steps for each high-risk patient.

For patients classified to be high-risk by ML model, social workers provided intensive assessment of psychosocial and quality of life barriers, and when appropriate, offered select high-risk patients additional psychosocial evaluations/interventions to target identified barriers through the Social Work Intensive (SWI) program. In the SWI, patient reported outcome questionnaires were administered before and after the intervention, and social workers charted additional notes on the encounters in the EMR. Dietitians utilized a high-risk assessment workflow looking at weight, nutrition, and access to food and supplements. Nurses assessed high-risk patients focusing on anemia, adequacy, dialysis access, blood pressure, fluid management, prior hospitalizations, glycemic control, and risk of skin ulcers and blood stream infection. Although additional interventions were intended to have been performed by social workers, nurses, and dietitians, the visits were charted in free text fields for routine notes and there was not unique frequencies of notes or distinct areas to chart interventions in the EMR.

### Social Worker Intensive Program

The SWI program was one of several components of the DHRP intervention. SWI was a previously established program throughout the provider’s national network of clinics, whereby social workers provided additional psychosocial evaluations/interventions (without the ML model) based on identified issues with treatment adherence, achievement of clinical quality targets, and/or other complications. This program demonstrated positive quality of life and hospitalization outcomes [17, 18].

In the DHRP, social workers used the ML model report to direct the SWI behavioral health interventions and dedicated additional time for all patients identified to be at a high risk of hospitalization. High-risk patients with psychosocial concerns were screened with a sleep quality survey (**Supplemental File 1**), psychological stress survey (**Supplemental File 2**) and the Centre for Epidemiological Studies Depression Scale (CES-D-10) survey [19], all of which are rated on a 1-10 Likert scale. Patients that screened positive for any barriers with sleep, distress, or depressive symptoms were eligible to participate in an 8-week SWI program with an optional 8-week extension (total of 16 weeks) for patients with continuing barriers in self-reported psychosocial outcomes. In the SWI program, the social workers delivered individually tailored cognitive, behavioral, sleep and interpersonal counseling to reduce any patient distress identified by the screening. At the end of the 8-week period, patients were re-screened with the surveys to assess intervention associated improvements in the outcomes. Social workers entered the pre and post screening measurements in the EMR.

### Statistical Methods

#### Descriptive Statistics

Descriptive statistics were tabulated for demographics and clinical parameters for patients in DHRP clinics and patients in control clinics.

#### Measuring Social Worker Intervention

We were unable to measure most DHRP related interventions performed by the interdisciplinary team members explicitly due to the limitations in the EMR documentation systems. However, we were able to use social worker evaluation notes and the psychosocial screeners administered in the SWI program as a measure of interventions performed for select high-risk patients. We used the number of assessment and intervention notes recorded by the social workers in the EMR to measure the SWI intervention before and after being identified as high-risk by the ML model. A two-sample T-statistic was used to evaluate the difference between the average number of EMR notes before and after being identified as high-risk. Similarly, T-statistics were used to see if there is a difference in the average depression, stress, and sleep scores before and after the SWI program.

#### Clinic Wide Hospital Admission and Hospitalization days rate

Data from patients at participating clinics were collected and clinic level rolling yearly hospital admission and hospital day rates per patient year (ppy) were calculated at baseline (2015) and 3 years after (2016-2018) the DHRP program started. Outcomes of the DHRP clinics were compared against control clinics that did not receive the high-risk patient report generated from the ML model. We computed the difference between annual hospital admission and day rates in DHRP and control clinics at baseline (2015) and at the end of follow up (2018). Paired t-tests were used to evaluate the change in the between group difference in the hospital admission and day rates from baseline (2015) to the end of the analysis period (2018).

## Results

### Clinic and Patient Characteristics

We used data from all active in-center HD patients in a DHRP and control clinics across the United States over the analysis period from 2016-2018. There were 54 DHRP clinics that had 7767 to 8189 active patients per year during the analysis period. There were 54 control clinics that had 7484 to 7705 active patients each year during the analysis period. The characteristics of patients at DHRP and control clinics is shown for all distinct patients in the clinic groups at baseline during 2015 (**Table 2**) and during the follow up period between 2016-2018 (**Table 3**).

**Table 3:**
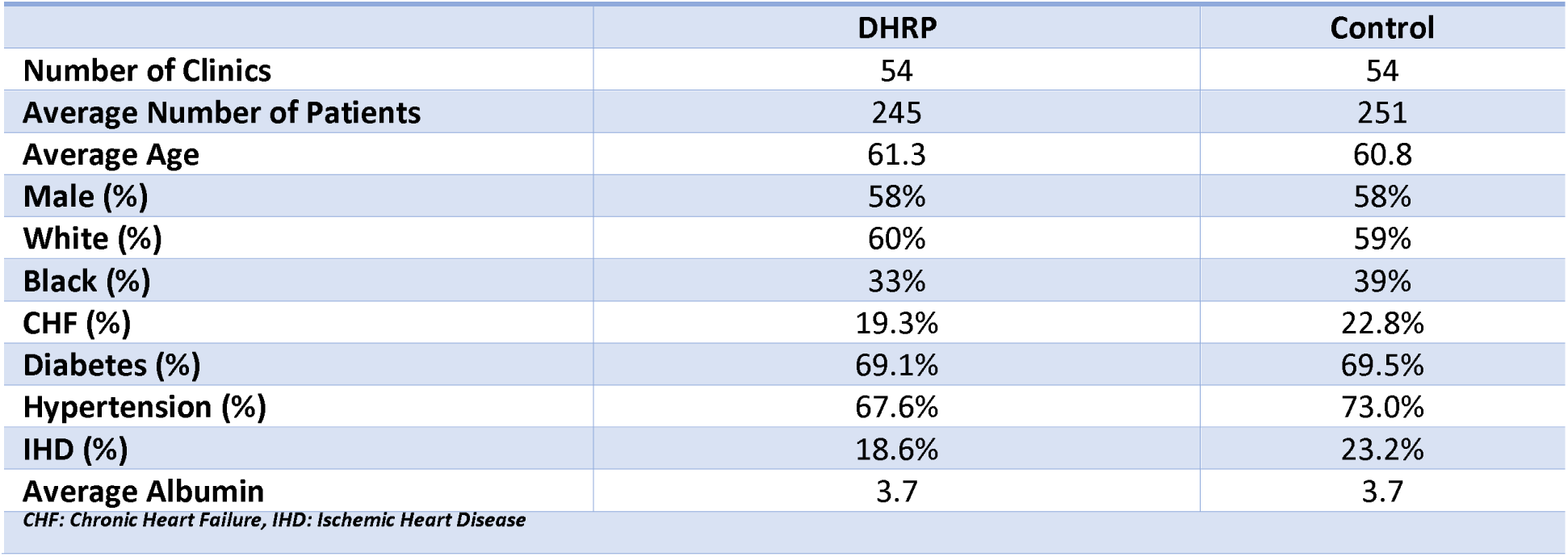
Demographic and comorbidities breakdown of patients in DHRP and control clinics during follow-up period 2016-2018.

During the analysis period, 1084 unique patients were identified as high-risk (predicted to have ≥6 hospital admissions in the following 12 months) by the ML model in the DHRP clinics. Within the DHRP clinics, the interdisciplinary teams evaluated and intervened on all high-risk patients. Fourteen percent (14%) of the patients (n=150) predicted to be at a high-risk by the ML model received the behavioral health screening by the social workers. The remaining ten percent (10%) of the total patients identified as high risk (n=111) were enrolled in the SWI program.

### Measuring Social Worker Intervention

Among the subset of high-risk patients identified who received behavioral health screening by the social workers (n=150), we found the average number of social work assessment and intervention notes charted in the EMR increased from 1.34±1.02 notes per month in 90 days before to 1.70±1.24 notes in the 90 days after the risk classification (p=0.01).

In the select subset of high-risk patients who received the SWI intervention to address a psychosocial barrier identified (n=111), we found improvements in patient reported outcomes for depressive symptoms, sleep quality, and psychological stress. We observed 62% of high-risk patients enrolled in the SWI program reported an reduction in their depressive symptoms as seen by CESD-10 scores, 64% patients reported an improvement in their sleep quality scores, and 56% patients reported a reduction in their psychological stress scores (**Table 4**). After the 8 week SWI intervention, the average CESD-10 scores decreased from 11.4 to 8.12 (CESD-10 scale from 0 to 60 with lower values representing fewer depressive symptoms), sleep scores decreased from 23.2 to 19.7 (sleep screener scale 5 to 50 with lower values representing better sleep quality), and stress scores decreased from 17.0 to 15.9 (stress screener scale 4 to 40 with lower values representing less psychological stress).

**Table 4:** Average Scores Before and After Social Worker Intervention.

| Survey | Number of Patients | Before Intervention Mean±SD (Range) | After Intervention Mean±SD (Range) | P-value |
| --- | --- | --- | --- | --- |
| CESD-10 Score | 81 | 11.42±6.17 (0-27) | 8.12 ±6.32 (0-21.5) | <0.0001 |
| Sleep Score | 72 | 23.22±10.69 (5-50) | 19.68 ±11.51 (5-49) | 0.0019 |
| Stress Score | 67 | 17.01±10.17 (1-40) | 15.85±8.28 (1-35) | 0.0055 |

### Clinic Wide Hospitalization rate

Annual rolling hospital admission rates had increasing trends over the analysis period in DHRP and control clinics. Cumulatively, the DHRP clinics exhibited a lower growth in admission and day rates as compared to control clinics (**Figure 3A and 3B**). At the end of follow up in 2018, the annual admission rate among DHRP clinics was 10% lower and the hospital day rate was 8% lower than control clinics. The DHRP clinics had a 5% and 2% increase in the hospital admission and day rates from 2015 to 2018, respectively. The control clinics showed a 15% and 10% increase in the hospital admission and day rates from 2015 to 2018, respectively.

**Figure 3A:**
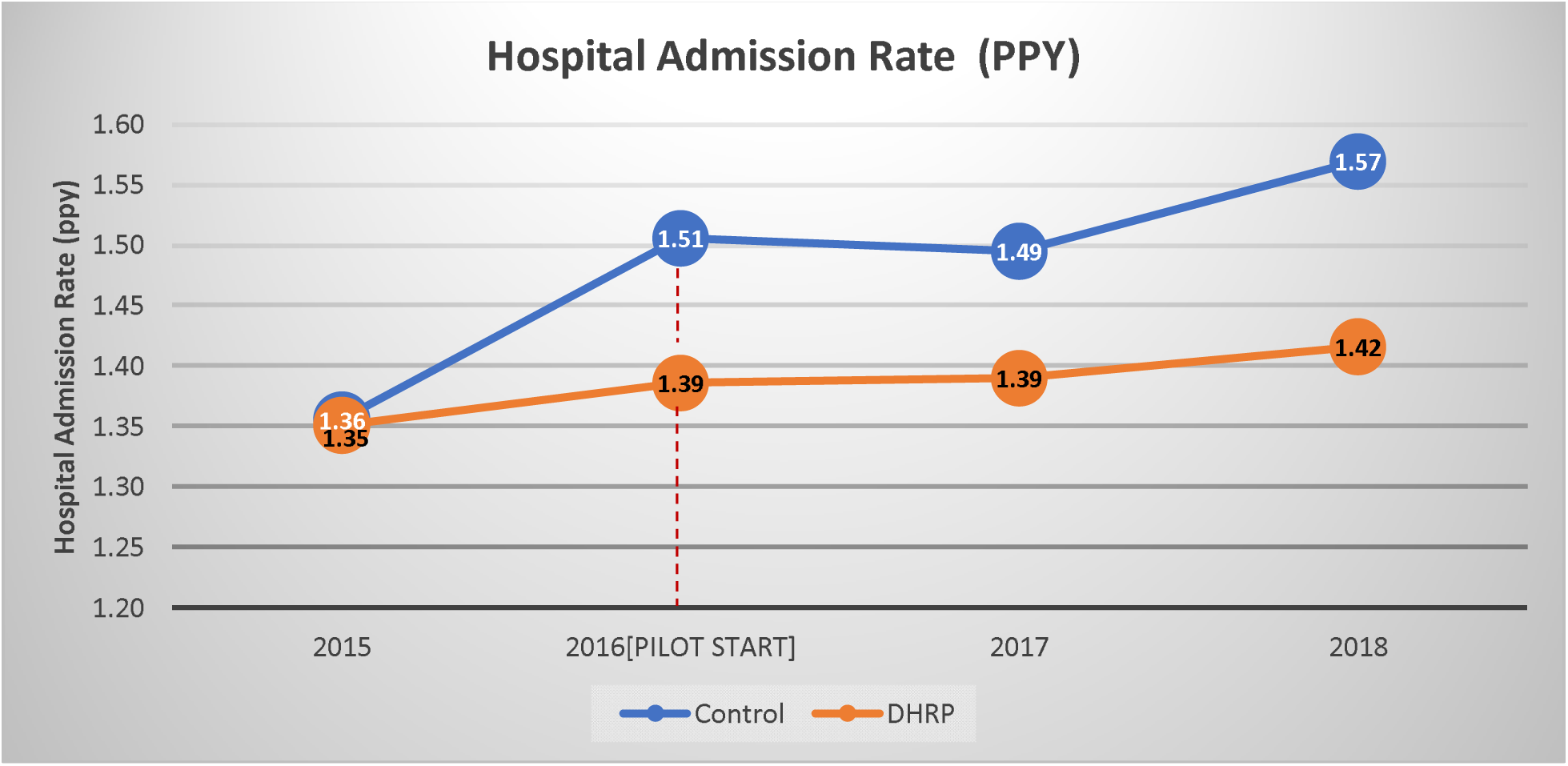
Hospitalization rate for DHRP clinics and control clinics at baseline and in follow-up period.

**Figure 3B:**
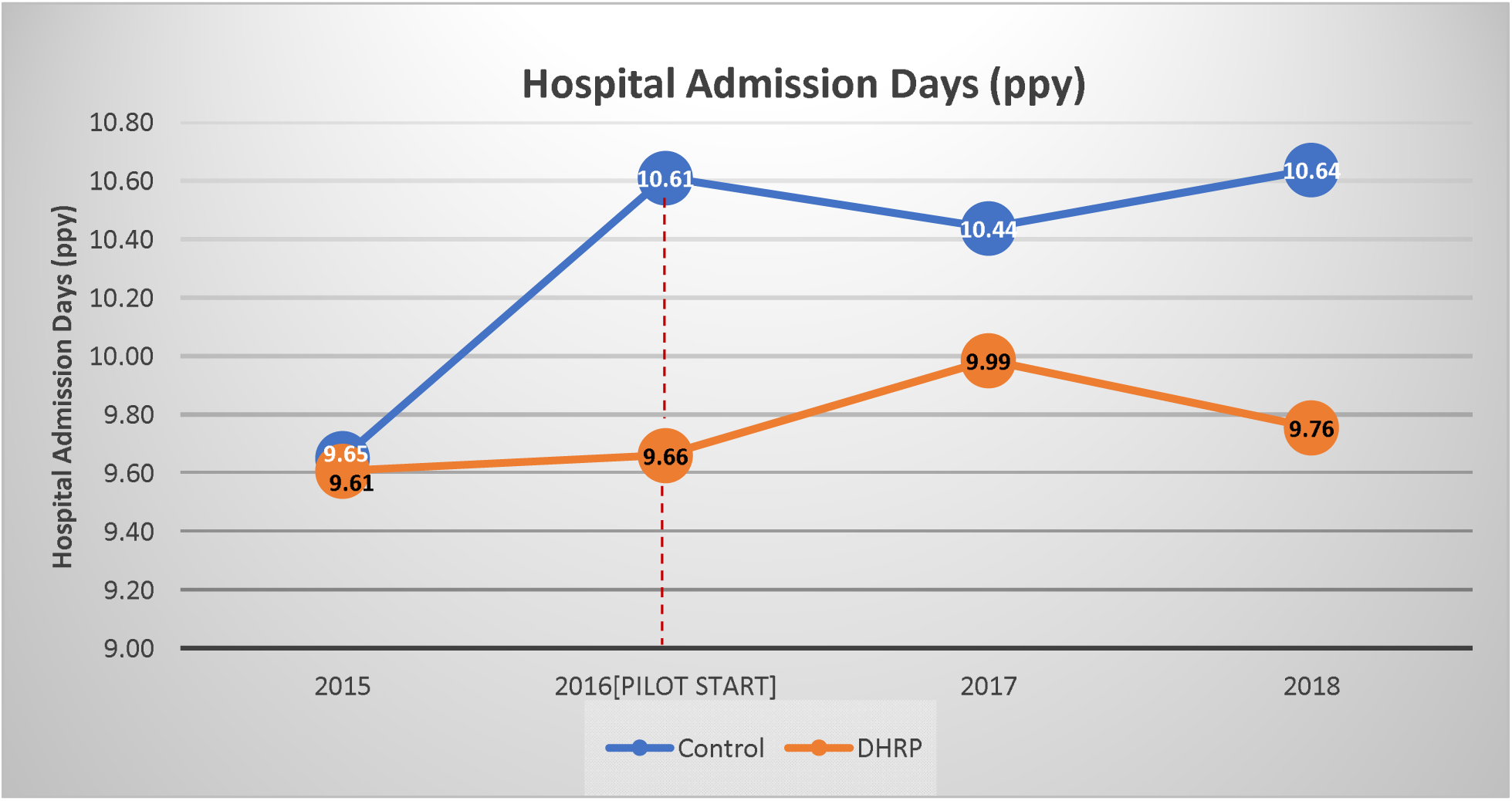
Hospitalization days rate for DHRP clinics and control clinics at baseline and in follow-up period.

The between group difference in rolling annual hospital admission and day rates was similar at baseline (2015) with a mean difference between DHRP versus control clinics of −0.008±0.09 ppy for the admission rate and −0.05±0.96 ppy for the hospital day rate. The between group difference in hospital admission and day rates became more distinct at the end of follow up (2018) with the mean difference in DHRP clinics being −0.155±0.38 ppy and −0.97±2.78 ppy lower than control clinics, respectively. A paired t-test showed the change in the between group difference in hospital admission and day rates from baseline to the end of the follow up was statistically significant (t-value=2.73, p-value<0.01 for admission rates; t-value=2.29, p-value=0.02 for hospital day rates).

## Discussion

We found ML directed care in outpatient dialysis clinics participating in the DHRP was associated with lower annual rolling all-cause hospitalization rates compared to matched control clinics. DHRP and control clinics exhibited consistent hospitalization rates at baseline in 2015. Although both groups of clinics had increases in hospitalization rates over time, the DHRP clinics had smaller increases compared to control clinics. At the end of follow up in 2018, the between group difference in hospitalization rates was distinct as compared to baseline and showed DHRP clinics had significantly lower hospital admission and day rates versus control clinics. In 2018, this distinction suggests that DHRP was potentially associated with 495 avoided hospital admissions and 2074 avoided hospital days as compared to matched control clinics. It appears monthly ML classifications for patients at a high-risk for multiple hospitalizations (≥6 admissions) in the next year helped direct interdisciplinary team interventions in the DHRP program and lessened the rising trends in the clinics’ hospitalization rates over the years. We found 14% of high risk patients were assessed for psychosocial barriers and social workers charted a higher number of notes 90 days after individual patients were identified to be at a high-risk for multiple hospitalizations. Among this subset of high-risk patients, 74% had a psychosocial barrier identified and received tailored behavioral health interventions from the social workers and we observed improvements in patient-reported outcomes for depressive symptoms, sleep quality, and psychological stress. Overall, it appears the ML model was able to aid the DHRP care teams in identifying the right individuals to target for personalized interventions at the right time, overall yielding improvements observable in outcomes at the clinic level.

The risk of hospitalization in ESKD patients is multifactorial and dependent on the etiology of the individual’s complication(s). Oftentimes the onset of a disease requiring inpatient care is marked by subtle temporal changes in patient presentations and clinical markers making it a challenge to universally classify hospitalization risk levels during standard of care dialysis practices. To the best of the authors’ knowledge, there are no other examples of AI/ML based hospitalization risk models being used to direct care in quality improvement efforts in dialysis. In general, the clinical application of AI in nephrology is scarce with only one report identified in a recent bibliometric study on the global evolution of AI in healthcare [20, 21]. The all-cause hospitalization risk models implemented in the DHRP appeared to suitably assist care teams with classifying the subset of individual patients in their clinic at the highest risk. Due to the nature of all-cause risk classification, effective root cause assessment methods to identify complications and target them with effective interventions was essential to yielding the positive results observed.

A recent review article on the factors and barriers related to decreasing admission rates in dialysis patients proposed three key clinical categories driving hospitalization risk that are potentially actionable [22]. These categories include volume control, infection, and psychosocial risks. The DHRP root cause assessments covered these categories, as well as other key areas such as nutrition, dialysis access, glycemic control, and ulcers. While most of these are fundamental areas of focus in dialysis care, psychosocial and behavioral health barriers are often not fully identified in standard care and span outside the field of nephrology [23, 24]. High risk patients, in fact, are often seen by the medical team as “too ill” to receive behavioral health intervention and, thus its value is forfeited. Depressive affect and sleep disturbances are known to associate with hospitalization rates and poor outcomes in chronic kidney disease patients [25-27]. The targeted interventions designed to improve mood, sleep quality and perceived stress appear to be possible contributors to the favorable hospitalization rates observed, albeit they were only performed in a small proportion of high-risk patients. Independent analyses are needed to understand the impacts of improvements for each patient reported measure, which include proprietary measures for sleep and psychological stress that have not been validated. Nonetheless, in general it appears the use of ML risk classifications for directing assessments and interventions selected by the patients’ care team may help personalize care and improve outcomes.

Although these findings on the application of ML directed care in the DHRP are of importance, there are some limitations to be considered. We only had data to define ML directed assessments specifically by social worker notes charted for concerns of psychosocial barriers, and sleep, depressive symptom, and stress scores were only available on a small subset of patients who received the SWI behavioral health intervention. Although we had some data on select social worker assessments/interventions, the EMR system of the provider was not structured in a manner that other ML directed assessments/interventions by social workers, dieticians, and nurses could be estimated. Nonetheless, the delivery of personalized interventions in line with needs, preferences, and goals of individuals adds to the novelty of the DHRP and is consistent with the paradigm shifts towards more patient centric care models [28, 29]. Having clinicians use their medical discretion to select interventions based on their assessment of each patient is fundamental in healthcare and appeared effective in this experience.

The successful application of ML directed care in the DHRP holds promise for further development and adoption of ML models that have the potential to provide an intelligent and timely triage of additional resources. Personalized interventions appear prudent to consider in ML directed endeavors, along with a toolbox of assessments and interventions that are anticipated to be effective in improving the quality of life and outcomes of patients. It may be important for the renal care industry to identify ways to increase early access to behavioral health care as patient’s increase their risk of hospitalization; using ML modeling might be helpful in directing patients toward this goal. However, ML based risk classification and other clinical decision support tools are never perfect. We believe it is of importance for clinical teams to understand the outputs generated and consider the limitations of the models in the design of directed assessments and interventions.

## Conclusions

We found ML directed assessment and personalized interventions in the DHRP, which included behavioral health intervention for some high-risk patients, were associated with lower all-cause hospitalization rates compared to control clinics. The DHRP efforts and findings detail an example of how the clinical application of AI directed care can be successfully conducted and will be of importance for considerations by payors, providers, and clinicians.

## Data Availability

The raw dataset and computer codes utilized for this analysis are not publically available information.

## General

A preliminary analysis of ML directed care in the DHRP was published as an abstract at the European Renal Association – European Dialysis Transplantation Association (ERA-EDTA) congress in 2020[30].

## Authors’ contributions

Design was performed by SC, HH, JL, YW, SJS, AV and LAU. Data Extraction and analysis was performed by SC, HH, JL and YW. The interpretation, drafting and revision of this manuscript was conducted by all authors. The decision to submit this manuscript for publication was jointly made by all authors and the manuscript was confirmed to be accurate and approved by all authors.

## Other contributions

We would like to acknowledge Vladimir M Rigodon for assistance with the composition of the regulatory protocol for this analysis.

---

**Supplemental File 1.**

|  |  |
| --- | --- |
| **Patient Name:** | **Date of Pre-Screening:** |
|  | **Date of Post Screening:** |

Sleep Screener
| Sleep problem identified | How bothered are you by it? Rate 1-10<br>(1=not at all)<br>(10 = very bothered) | Sleep problem identified | How bothered are you by it? Rate 1-10<br>(1=not at all)<br>(10 = very bothered) |
| --- | --- | --- | --- |
| 1. Difficulty *falling* asleep | 1 2 3 4 5<br>6 7 8 9 10 | 4. *Interrupted* sleep<br>(awakening, then falling asleep again on and off through the night) | 1 2 3 4 5<br>6 7 8 9 10 |
| 2. Difficulty *staying* asleep | 1 2 3 4 5<br>6 7 8 9 10 | 5. *Restless legs* that awaken you<br>*Restless legs syndrome is the uncomfortable feeling in legs or an urge to keep moving your legs | 1 2 3 4 5<br>6 7 8 9 10 |
| 3. Difficulty *awakening* | 1 2 3 4 5<br>6 7 8 9 10 |  |  |

| Name | Name of Medication | Taken for what symptoms? | How often do you take them? |
| --- | --- | --- | --- |

**Has patient ever been referred for a sleep evaluation?** ☐Yes ☐No
**Has patient ever participated in a sleep evaluation?** ☐Yes ☐No
Comments:

**Supplemental File 2.**
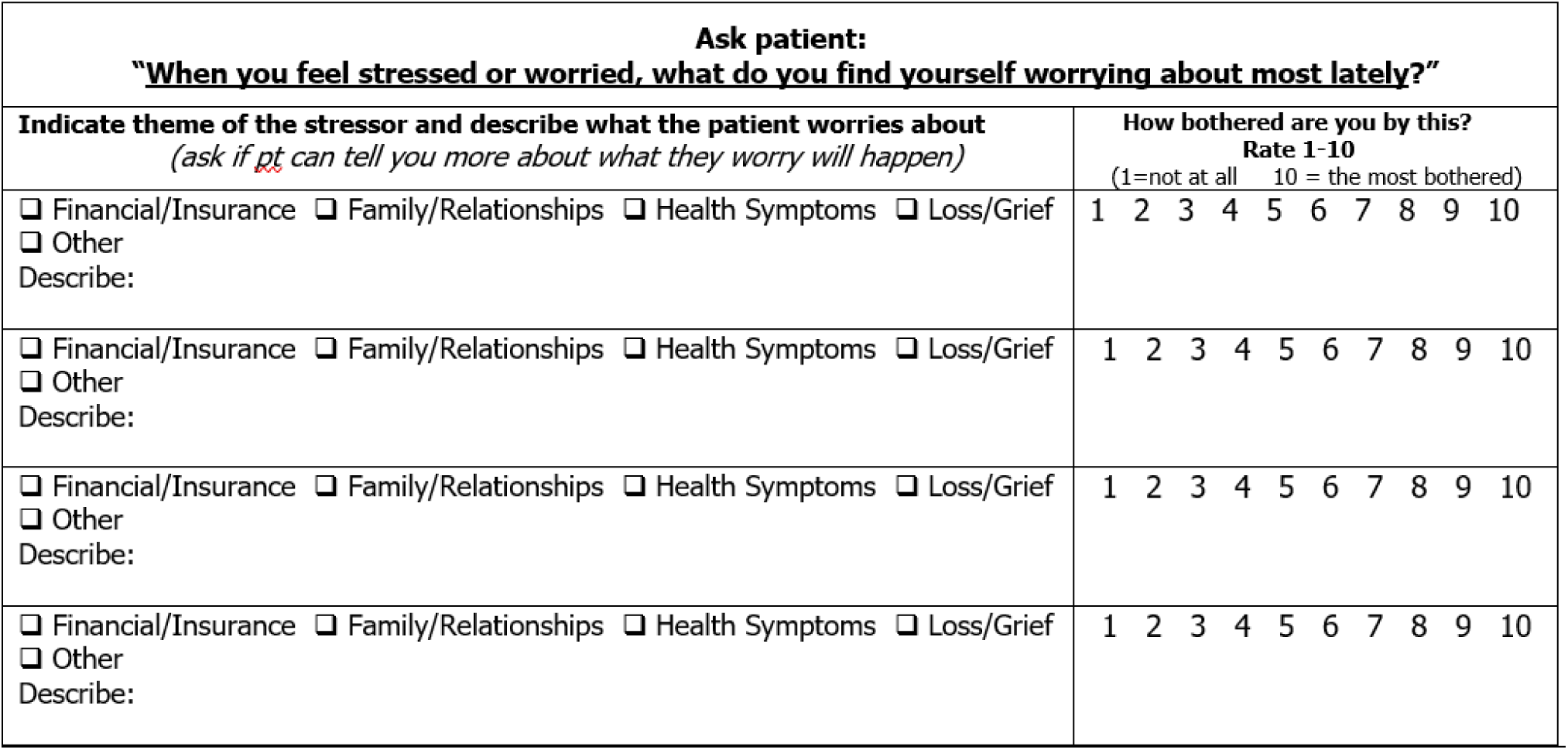

